# Dissociable computational markers of semantic search and verbal retrieval drive across the psychosis spectrum

**DOI:** 10.64898/2026.05.18.26353478

**Authors:** Roya M. Hüppi, Werner Surbeck, Yves L. Pauli, Noemi Dannecker, Dominic Fabian, Victoria Edkins, Sandra A. Just, Niklaus Denier, Tobias Bracht, Frederike Stein, Rieke R. Mülfarth, Svenja Seuffert, Tilo Kircher, Iris E. Sommer, Wolfram Hinzen, Philipp Homan

## Abstract

Formal thought disorder (FTD) is a core psychosis feature. Disentangling its dimensions requires tasks simple enough for formal modeling yet sensitive enough to capture individual variation across the psychosis spectrum. The semantic verbal fluency task offers precisely this: a structured behavioral trace of semantic memory sampling, amenable to computational analysis using distributed word embeddings. We hypothesized that this sampling process is governed by two dissociable mechanisms mapping onto FTD dimensions: initial retrieval drive (*d*_0_), quantifying the motivational resource sustaining production, and semantic search precision (*α*), quantifying how strongly similarity to the preceding word constrains each retrieval step from near-random to highly structured. We hypothesized that reduced *d*_0_ would track negative psychosis symptoms and alogia, while degraded *α* would track language disorganization and left inferior longitudinal fasciculus (ILF) fractional anisotropy. We tested these predictions in a primary (*N* = 120) and an independent replication sample (*N* = 249) of German-speaking individuals across the psychosis spectrum. Both parameters decreased with greater psychosis severity and, in the primary sample, they dissociated regarding their clinical correlates. *d*_0_ correlated negatively with negative symptoms, general psychopathology, and poverty of speech, consistent with a computational signature of alogia. *α* correlated negatively with positive symptoms and cognitive flexibility, and, in individuals with psychosis, positively with left ILF fractional anisotropy. The association between *d*_0_ and negative symptoms was replicated in the independent sample. These findings pave the way for mechanistic, automatically derived FTD markers capturing subclinical variation across the psychosis spectrum and mapping onto underlying cognitive and neural processes.

## Introduction

Formal thought disorder (FTD) is observable as incoherent, disorganized, or impoverished language. It is among the most clinically consequential features of psychosis, correlating with functional outcomes and predicting relapse [1–3]. Its mechanistic basis, however, remains opaque. Negative FTD, characterized by poverty of speech and content, is more pronounced in schizophrenia than in affective psychoses and often persists in remission in individuals with schizophrenia. Positive FTD, characterized by derailment and incoherence, occurs across diagnostic boundaries and remits with treatment in affective psychoses, yet often persists in stable individuals with schizophrenia [1, 4]. This clinical dissociation suggests at least two partially independent processes underlying language abnormalities in psychosis: one relates to the individual’s verbal retrieval drive and the other one captures how the semantic space is explored. Since most signs of FTD may be latent and present as subtle degradations of semantic structure that, even with proper training, fall below the threshold of clinical detection, a computational approach is warranted. By automatically and continuously quantifying the degree of semantic constraint in language production, variations that are too subtle for clinical ratings to capture reliably become accessible [5–10]. This enables new applications in clinical trials aiming to identify and prevent clinical worsening based on language [11].

The easily administered semantic verbal fluency task offers a particularly tractable window onto individual differences in retrieval drive and the navigation of semantic space during language production. Asked, for example, to name as many animals as possible in sixty seconds [12], speakers repeatedly sample from semantic memory, and the produced sequence of words reveals their sampling structure [13]. Computational analysis of pairwise semantic distances between consecutive responses, made possible by distributed word embeddings [14, 15], shows that healthy speakers traverse semantic space in a locally coherent manner, clustering semantically related items before transitioning to new regions [13, 16]. Individuals with schizophrenia produce shorter lists and show reduced semantic similarity between consecutive responses compared to healthy individuals[17–19], with lower semantic similarity being demonstrated by individuals with signs of FTD than by those without markers of FTD [20, 21]. Foraging models provide a principled account of the semantic search in verbal fluency tasks: retrieval resembles optimal foraging in patchy environments, with local exploitation of semantic clusters interspersed with global transitions [16, 22]. Recent work applying this framework to psychosis showed that the degree to which semantic similarity constrains word selection is reduced in schizophrenia and tracks negative symptom severity [23]. We build on this by asking a more specific question: not just whether semantic guidance is reduced, but what drives that reduction and whether it dissociates from the motivational capacity to sustain retrieval at all. We model each participant’s fluency list as the output of a retrieval process governed by two parameters (Figure 1). Initial retrieval drive (*d*_0_), captures the motivational resource sustaining word production, which closely tracks list length. Semantic search precision (*α*) captures how strongly similarity constrains the exploration of semantic space in each sampling step. When *α* is high, retrieval is structured and locally coherent. When *α* approaches zero, retrieval approaches a random walk through semantic space, indifferent to the similarity structure of memory. These parameters are formally orthogonal: *d*_0_ is identified by the length of the list, *α* by its transition structure, and they are estimated in a two-stage procedure that prevents one from absorbing variance belonging to the other.

**Figure 1:**
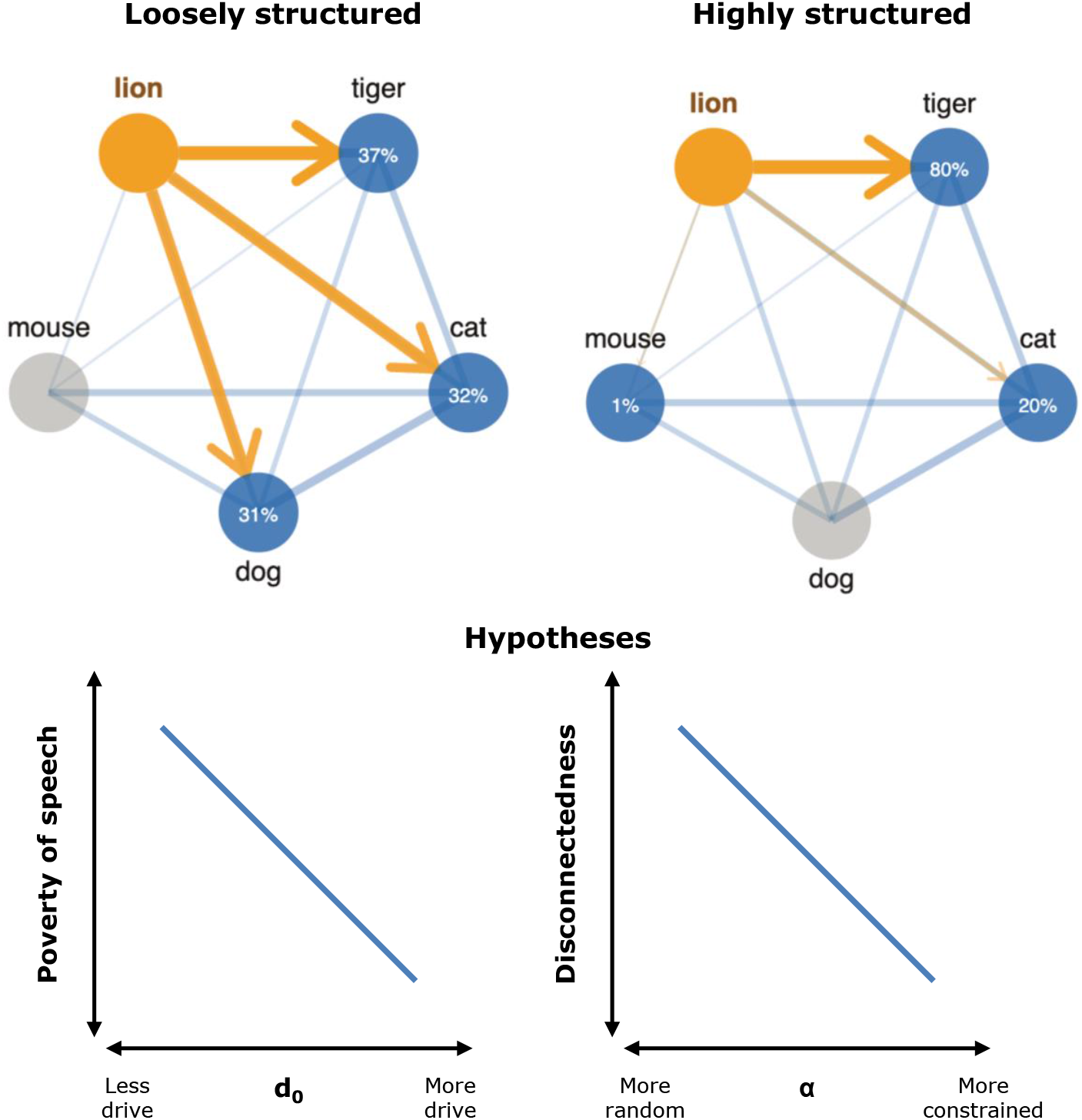
Model scheme. *Top*: Two example retrievals from a five-item semantic network starting from lion, illustrating the effect of semantic search precision (*α*) on the sampling process. Edge thickness reflects semantic similarity. In the loosely structured case (low *α*, left), probabilities are distributed more evenly across candidates regardless of similarity, and the walk jumps with low predictability through semantic space, a pattern consistent with the loosened associations observed in positive formal thought disorder. In the highly structured case (high *α*, right), the selection probabilities are strongly concentrated on the most similar unvisited item: tiger receives 80% of the probability mass at the first step, and the walk stays locally coherent throughout. *Bottom*: Hypothesized relationships between the two formally orthogonal model parameters and their clinical correlates. Initial retrieval drive (*d*_0_) is predicted to track the quantity of verbal output, with lower *d*_0_ reflecting less motivational resource sustaining production before stopping and thus higher poverty of speech. *α* is predicted to track language disorganization, with lower *α* reflecting a more random walk through semantic space and thus higher disconnectedness in speech.

The dissociation between *d*_0_ and *α* carries theoretical hypotheses. We hypothesized that lower *d*_0_ is associated with higher negative psychosis symptom burden and higher clinical ratings of poverty of speech, rather than with language disorganization per se [24, 25], constituting a computational proxy for alogia. For *α*, we hypothesized that lower *α* is associated with higher positive psychosis symptom burden and higher clinical ratings of disorganized speech [26, 27]. Additionally, we hypothesized that *α* correlates positively with cognitive flexibility, since structured semantic search is cognitively demanding [13], as well as fractional anisotropy of the ventral language stream, particularly the inferior longitudinal fasciculus, which supports semantic representation [28, 29]. We tested these hypotheses across two independent samples spanning the psychosis spectrum, with the examination of the association between *α* and white matter microstructure of the ventral language stream being restricted to the primary sample.

## Methods and materials

### Primary sample – VELAS

#### Participants

Data were drawn from the VELAS study (“Ventral language stream in schizophrenia with regard to semantic and visuo-spatial processing anomalies”), conducted at the University Hospital of Psychiatry Zurich in Zurich, Switzerland [30–32]. The sample comprised *N* = 120 participants across three groups: individuals with early psychosis (*n* = 40), individuals with high schizotypy (*n* = 40), and individuals with low schizotypy (*n* = 40). Individuals with psychosis fulfilled the ICD-10 criteria for a psychotic disorder (schizophrenia (F20; *n* = 22), acute and transient psychotic disorder (F23; *n* = 10), schizoaffective disorder (F25; *n* = 5), recurrent depressive disorder, current episode severe with psychotic symptoms (F33.3; *n* = 3)) and they had to be within the first 8 years after the onset of psychosis. Categorization for the high and low schizotypy groups was determined via cluster analysis of scores from the Multidimensional Schizotypy Scale (MSS, [33]) and the Oxford-Liverpool Inventory of Feelings and Experiences (O-LIFE, [34]), derived through an online survey (*N* = 1061). Individuals clustering with the psychosis group were assigned to the high schizotypy group, while the low schizotypy group was drawn from clusters with the lowest scores. Both groups were demographically matched to the psychosis group in terms of sex, educational level, and age. All participants had to be fluent in German. Exclusion criteria included neurological disorders and substance intoxication or withdrawal in the last month. Written informed consent was obtained from all participants. The study was approved by the Cantonal Ethics Committee Zurich (Kantonale Ethikkommission Zürich; Protocol No. 2021-00347).

#### Clinical measures

In the VELAS sample, psychopathology was assessed with the Positive and Negative Syndrome Scale (PANSS) [35] and schizotypal traits with the MSS [33] and the O-LIFE [34]. FTD was evaluated with the Scale for the Assessment of Thought, Language, and Communication (TLC) [1]. Within the TLC, verbal underproductivity and disconnected speech have previously been identified as two distinct subtypes of communication abnormalities using confirmatory factor analysis. Verbal underproductivity is measured using the poverty of speech item from the TLC. Disconnected speech is defined as the mean of five TLC items: tangentiality, derailment, incoherence, circumstantiality, and loss of goal [36]. Cognitive flexibility was indexed by the Trail Making Test B minus A score (TMT B−A) [37], where higher scores indicate lower cognitive flexibility, reflecting the additional time cost in seconds of set-shifting relative to simple sequencing. The IQ score from the Mehrfachwahl-Wortschatz-Intelligenztest (MWT-B) [38] served as a proxy for premorbid IQ.

#### Neuroimaging

Imaging was conducted on a Philips Achieva 3.0 T magnetic resonance imaging scanner using a 32-channel SENSE head coil (Philips, Best, The Netherlands). The protocol included 3D-T1-weighted imaging with a voxel resolution of 1 mm^3^ and diffusion weighted imaging (DWI) with 64 non-collinear directions (3000 s/mm^2^) and one b0 (0 s/mm^2^), with a voxel resolution of 1.96 × 1.96 × 2 mm^3^. Details on DWI data processing have previously been described [31].

##### Manual tractography

Manual tractography was performed using region-of-interest (ROI) masks to reconstruct the inferior longitudinal fasciculus (ILF), a main pathway of the ventral language stream [39] (Figure 4A). One ROI was positioned within the white matter of the anterior temporal lobe, just anterior to the temporal horn of the ventricular system, and one in the coronal plane at the level of the preoccipital notch [40]. Fibers that belonged to neighboring fiber systems were manually removed using carefully placed regions of avoidance. Fractional anisotropy (FA) values were extracted for the described tracts using tract-based spatial statistics and deterministic tractography.

### Replication sample – FOR2107

#### Participants

The replication sample was drawn from the FOR2107 consortium study conducted in Marburg, Germany [41]. 264 participants were initially enrolled. Of these, 262 had available fluency data. 12 participants were subsequently excluded due to missing data within the fluency task, and 1 additional participant was excluded due to missing demographic and clinical information. The final sample therefore consisted of *N* = 249 participants in three groups that were analogous with those in the VELAS sample: individuals with a DSM-IV diagnosis of schizophrenia (*n* = 83), individuals with high schizotypy (*n* = 84), and individuals with low schizotypy (*n* = 82). Schizotypy group membership was determined by score on the Schizotypal Personality Questionnaire (SPQ-B) [42], with high and low schizotypy defined by empirically derived cut-offs (i.e., a SPQ-B sum score of *>* 3 vs. ≤ 3). All participants gave written informed consent. The FOR2107 study was approved by the Ethics Committees of the Medical Faculties, University of Marburg (AZ: 07/14) and University of Münster (AZ: 2014-422-b-S).

#### Clinical measures

In the FOR2107 sample, the Scale for the Assessment of Negative Symptoms (SANS) [43] and the Scale for the Assessment of Positive Symptoms (SAPS) [44] were used to assess psychopathology. Schizotypal traits were evaluated with the SPQ-B [42]. The MWT-B IQ score served as a proxy for premorbid IQ [38]. FTD and cognitive flexibility were not assessed in this sample.

#### Verbal fluency task and semantic space construction

Participants were asked to name as many animals as possible within 60 seconds. Responses were audio-recorded and transcribed. Prior to analysis, transcripts were cleaned using a standardized replacement dictionary. This covered four categories of corrections: foreign-language responses mapped to their German equivalents (e.g., *Buffalo, Büffel*), Swiss German dialect variants mapped to standard German (e.g., *Geiss, Ziege*), typographic errors corrected (e.g., *Haase, Hase*), and plural forms reduced to their singular canonical form (e.g., *Wölfe, Wolf*). A small number of items were excluded entirely prior to vocabulary construction: words for which the fastText representation was insufficiently connected to the animal semantic space, because the word was too rare in general German text, or because the dominant meaning of the word in the training corpus was not the animal meaning (e.g., *Bremse*, which primarily refers to a brake rather than a horsefly). A shared vocabulary was then constructed as the union of all items produced across all participants. Word vectors were obtained from a pre-trained German fastText model [14, 15] trained on Common Crawl and Wikipedia (300-dimensional, subword skipgram) using an in-house developed modular Python package called pelican_nlp (https://github.com/ypauli/pelicannlp) [45]. For each vocabulary item *i*, its embedding **v**_*i*_ ∈ R^300^ was retrieved, and pairwise cosine similarities were computed to form the similarity matrix *S* ∈ R^*V* ×*V*^, where *V* is the vocabulary size:

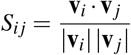

The diagonal *S*_*ii*_ = 1 by construction. All subsequent model computations operate on this fixed similarity matrix.

#### Computational model of semantic verbal fluency

We model verbal fluency production as a foraging process on a weighted semantic graph [16, 22]. At each step *t*, the speaker selects the next word from a neighborhood of unvisited candidates, guided by semantic similarity to the most recently produced word. Production terminates when motivational drive falls below a threshold determined probabilistically. Specifically, at step *t*, the neighborhood *N*_*t*_ is the set of all vocabulary items not yet produced. If a similarity threshold *τ* is applied, *N*_*t*_ is restricted to items with *S*_*j*,prev_ ≥ *τ*, where prev denotes the most recently produced word. If no connected neighbors remain, the full unvisited set is used as a fallback. Selection probabilities are based on rank rather than raw similarity values. This is analogous to using Spearman rather than Pearson correlation: rank order is preserved while sensitivity to the absolute scale of the embedding space is eliminated. Since word embedding models trained on general corpora compress the similarity range within semantic categories [14], using raw values would attenuate the selection gradient in a way that confounds embedding quality with individual differences in search precision. The rank-based formulation is robust to this compression.

Hence, for the model, words are selected with probability proportional to a rank-weighted distribution. Let *r*_*j*_ denote the rank of item *j* in *N*_*t*_ when sorted by *S*_*j*,prev_ in descending order (rank 1 = most similar). The selection probability is:

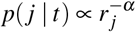

normalized over *𝒩*_*t*_, with the Zipf normalization constant 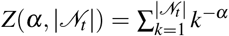. When *α* = 0, our suggested “surfer” model reduces to a random walk model [22]. When *α →* ∞, the most similar unvisited item is always selected. Free parameters are *d*_0_ and *α*.

#### Stopping mechanism

Motivational drive decays exponentially over the course of the list:

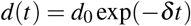

where *d*_0_ ∈ (0, 1) is the initial retrieval drive and *δ >* 0 is a fixed decay rate (see Calibration below). The probability of stopping at step *t* is:

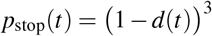

The cubic exponent ensures a gradual onset of stopping as drive depletes, consistent with empirical list-length distributions. On each step, the speaker stops with probability *p*_stop_(*t*); otherwise, a word is selected from the set of unvisited candidates.

#### Parameter estimation and two-stage M-step

Parameters were estimated by Expectation-Maximization (EM) [46], treating the latent state sequence (exploit, switch, stop) as the missing data. An empirical Bayes prior on the unconstrained parameters was updated at each iteration from the group posterior, regularizing estimates for participants with short lists. A key challenge is that *d*_0_ and *α* carry orthogonal signals in the data: *d*_0_ is identified by list length (the stopping trajectory), while *α* is identified by the rank structure of consecutive transitions (the emission trajectory). Jointly optimizing both parameters allows the emission term, which contributes ∼115 nats per list, to dominate the stopping term (∼3 nats), destroying identifiability of *d*_0_.

We resolved this with a two-stage M-step that exploits the orthogonality:

- **Stage 1**. *α* is updated from the emission log-likelihood only, holding *d*_0_ fixed. This likelihood depends on the rank of each observed word among its unvisited neighbors and is independent of *d*_0_.
- **Stage 2**. *d*_0_ is updated from the stopping log-likelihood only, holding *α* fixed. This likelihood depends on the sequence of survival and stopping probabilities and is independent of *α*.

#### Calibration of fixed parameters

The threshold *τ* was chosen to yield a mean neighborhood size of 200–250 connected words per vocabulary item, ensuring a high rate of connectedness. The decay rate *δ* was chosen so that the model’s simulated mean list length (under *d*_0_ = 0.70 and *α* = 1.0) matched the observed category mean. Calibrated values are reported in Table S1. For all three categories, calibration yielded *δ* = 0.003 and *τ* = 0.0.

#### Parameter recovery

To validate identifiability, we conducted a parameter recovery simulation using a synthetic block-diagonal similarity matrix (*V* = 60 items, 6 clusters of 10, within-cluster similarity = 0.70, between-cluster similarity = 0.20). We generated *N* = 50 synthetic participants with known *d*_0_ ∼ *𝒰* (0.35, 0.92) and *α* ∼ *𝒰* (0.30, 2.00), simulated fluency lists from the surfer model and refitted the model using the same EM procedure. Recovery was quantified as the Pearson correlation between true and estimated parameters; scatter plots of true versus recovered values, the joint log-likelihood surface, and examples of simulated lists under contrasting parameter regimes are shown in Figures S1, S2, and S3 in the Supplement.

### Statistical analysis

Group comparisons within datasets were performed using one-way ANOVA with Tukey’s HSD post-hoc tests for continuous variables and Pearson’s Chi-square test of independence for categorical variables. Imaging correlations were conducted in the full VELAS sample and, additionally, in the psychosis group only. All correlations were Pearson’s *r* unless otherwise noted. Comparability between the VELAS and FOR2107 cohorts was assessed for demographic characteristics. For continuous variables, equality of variance was first evaluated using Levene’s test. Since the assumption of homoscedasticity was not met, Welch’s *t*-test was used to account for unequal variances. Categorical variables were analyzed using Pearson’s Chi-square tests of independence. All statistical tests were two-tailed with the alpha level set at .05. All analyses were conducted in R and Python; code will be made available at https://github.com/homanlab/.

## Results

### Primary sample – VELAS

#### Sample characteristics

The VELAS sample comprised 120 participants (*n* = 40 in the low schizotypy, *n* = 40 in the high schizotypy, and *n* = 40 in the early psychosis group). Groups did not significantly differ regarding age (*F*(2, 117) = 2.94, *p* = .057), sex (*χ*^2^(2) = 5.34, *p* = .069), or premorbid IQ (MWT-B IQ: *F*(2, 117) = 2.74, *p* = .069). Sample characteristics are reported in Table 1.

**Table 1:**
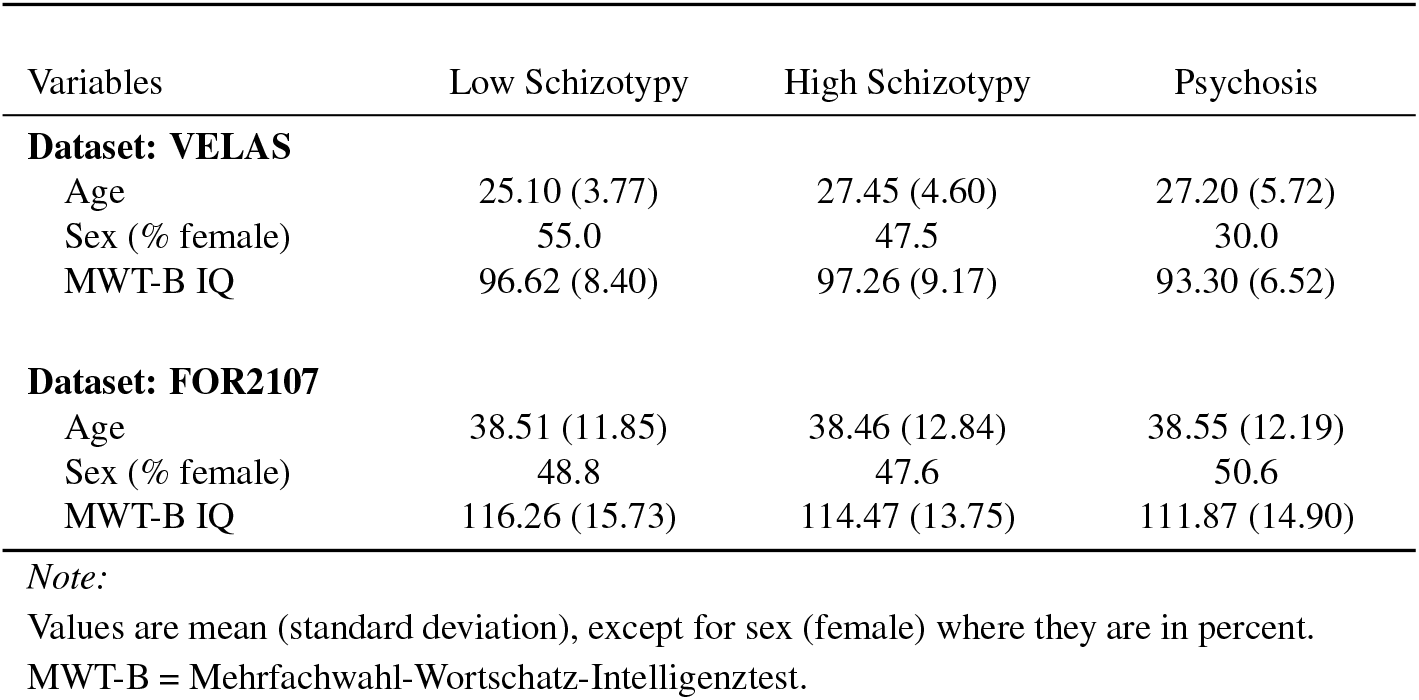
Sample characteristics in VELAS and FOR2107.

| Variables | Low Schizotypy | High Schizotypy | Psychosis |
| --- | --- | --- | --- |
| **Dataset: VELAS** |  |  |  |
| Age | 25.10 (3.77) | 27.45 (4.60) | 27.20 (5.72) |
| Sex (% female) | 55.0 | 47.5 | 30.0 |
| MWT-B IQ | 96.62 (8.40) | 97.26 (9.17) | 93.30 (6.52) |
| **Dataset: FOR2107** |  |  |  |
| Age | 38.51 (11.85) | 38.46 (12.84) | 38.55 (12.19) |
| Sex (% female) | 48.8 | 47.6 | 50.6 |
| MWT-B IQ | 116.26 (15.73) | 114.47 (13.75) | 111.87 (14.90) |

#### Clinical measures

A one-way ANOVA revealed significant group differences in the number of items produced in the verbal fluency task (*F*(2, 117) = 5.42, *p* = .006), with post-hoc Tukey’s HSD tests indicating that individuals with psychosis generated significantly less items than those with low schizotypy (*p* = .005). There was a significant main effect of group on the total PANSS scores (*F*(2, 117) = 132.90, *p* < .001), with the psychosis group showing significantly higher symptom burden than the low and high schizotypy group (*p* < .001 for each comparison). There were also significant group differences in overall FTD during conversational speech (TLC total: *F*(2, 117) = 22.04, *p* < .001), with individuals with psychosis showing more overall FTD than those with low and high schizotypy (*p* = < .001 for each comparison). Furthermore, there was a significant difference in cognitive flexibility between the groups (TMT B−A: *F*(2, 117) = 4.22, *p* = .017). Individuals with psychosis demonstrated significantly lower cognitive flexibility than individuals with low (*p* = .046) and high schizotypy (*p* = .027). An overview of the descriptive statistics and group comparisons is provided in Table 2.

**Table 2:** Clinical, Neural, and Model Parameters in VELAS and FOR2107.

| Variables | Low<br>Schizotypy<br>(LS) | High<br>Schizotypy<br>(HS) | Psychosis (P) | F-statistic | *p*-value | Post-hoc analyses <sup>a</sup> |  |  |
| --- | --- | --- | --- | --- | --- | --- | --- | --- |
|  |  |  |  |  |  | LS vs HS | LS vs P | HS vs P |
| **Dataset: VELAS** |  |  |  |  |  |  |  |  |
| Verbal fluency (animals) | 25.98 (6.15) | 24.65 (7.47) | 21.25 (6.16) | 5.42 | .006** | — | .005** | — |
| PANSS total | 32.12 (5.36) | 32.75 (3.66) | 61.73 (14.70) | 132.90 | < .001*** | — | < .001*** | < .001*** |
| PANSS positive | 7.40 (1.60) | 8.15 (1.49) | 14.18 (5.61) | 45.64 | < .001*** | — | < .001*** | < .001*** |
| PANSS negative | 8.10 (1.75) | 7.58 (0.81) | 17.07 (6.07) | 84.34 | < .001*** | — | < .001*** | < .001*** |
| PANSS general | 16.62 (2.77) | 17.02 (2.42) | 30.48 (6.75) | 126.11 | < .001*** | — | < .001*** | < .001*** |
| TLC total | 0.15 (0.53) | 0.13 (0.34) | 0.95 (0.87) | 22.04 | < .001*** | — | < .001*** | < .001*** |
| TLC poverty of speech | 0.00 (0.00) | 0.00 (0.00) | 0.50 (0.95) | 10.91 | < .001*** | — | < .001*** | < .001*** |
| TLC disconnectedness | 0.01 (0.09) | 0.08 (0.24) | 0.22 (0.48) | 4.23 | .017* | — | .014* | — |
| TMT B–A | 28.23 (14.15) | 29.08 (15.33) | 38.73 (22.79) | 4.22 | .017* | — | .027* | .046* |
| FA of left ILF | 0.43 (0.02) | 0.43 (0.03) | 0.43 (0.03) | 0.25 | .778 | — | — | — |
| *d*<sub>0</sub> | 0.70 (0.03) | 0.69 (0.03) | 0.67 (0.04) | 5.43 | .006** | — | .005** | — |
| *α* | 0.63 (0.22) | 0.63 (0.22) | 0.57 (0.24) | 1.00 | .372 | — | — | — |
| **Dataset: FOR2107** |  |  |  |  |  |  |  |  |
| Verbal fluency (animals) | 27.00 (6.06) | 25.63 (6.16) | 21.17 (5.31) | 22.43 | < .001*** | — | < .001*** | < .001*** |
| SAPS total | 0.17 (0.62) | 0.39 (1.28) | 8.87 (11.61) | 44.75 | < .001*** | — | < .001*** | < .001*** |
| SANS total | 0.54 (2.01) | 1.68 (3.93) | 14.06 (11.25) | 95.74 | < .001*** | — | < .001*** | < .001*** |
| SPQ-B total | 1.57 (1.19) | 6.92 (2.82) | 10.55 (5.16) | 139.97 | < .001*** | < .001*** | < .001*** | < .001*** |
| *d*<sub>0</sub> | 0.70 (0.03) | 0.69 (0.03) | 0.67 (0.03) | 20.27 | < .001*** | — | < .001*** | < .001*** |
| *α* | 0.62 (0.23) | 0.58 (0.21) | 0.61 (0.23) | 0.53 | .588 | — | — | — |

*Note:*
Values are mean (standard deviation). Group differences were tested with one-way ANOVA followed by Tukey's HSD post-hoc tests.
PANSS = Positive and Negative Syndrome Scale; TLC = Scale for the Assessment of Thought, Language, and Communication; TMT B–A = Trail Making Test B minus A; FA = fractional anisotropy; ILF = inferior longitudinal fasciculus; SAPS = Scale for the Assessment of Positive Symptoms; SANS = Scale for the Assessment of Negative Symptoms. SPQ-B = Schizotypal Personality Questionnaire.
\* $p < .05$ . \*\* $p < .01$ . \*\*\* $p < .001$ . <sup>a</sup> Only significant $p$ -values are reported.

### Model parameters

#### Parameter dissociation

*d*_0_ differed significantly between groups (*F*(2, 117) = 5.43, *p* = .006), with *d*_0_ being significantly lower in the psychosis group than the low schizotypy group (*p* = .005). It showed a strong correlation with the number of items produced (r = 0.95, *p* < .001), confirming that *d*_0_ captures the list-length signal as expected. Semantic search precision, *α*, did not differ significantly between groups. However, *α* was descriptively lower in individuals with psychosis (*M* = 0.57, *SD* = 0.24) than in individuals with low (*M* = 0.63, *SD* = 0.22) and high schizotypy (*M* = 0.63, *SD* = 0.22). It correlated significantly, although modestly, with the semantic similarity of consecutive items (r = 0.21, *p* = .019). The two parameters *d*_0_ and *α* were positively correlated (r = 0.23, *p* = .012), reflecting the general tendency for participants who produce longer lists to also show more structured semantic search (see Figure S4 in the Supplement). The association was modest, consistent with the two-stage estimation procedure which fits *d*_0_ and *α* from orthogonal signals (list length and transition structure respectively) such that the two parameters capture dissociable aspects of fluency production and are examined separately below. Table 2 provides an overview of the descriptive statistics and group comparisons.

#### Clinical associations

*d*_0_ was negatively associated with both total symptom burden (PANSS total: r = -0.38, *p* < .001) and negative symptom severity specifically (PANSS negative: r = -0.38, *p* < .001) across the full sample. Further, *d*_0_ was negatively correlated with poverty of speech during conversational speech (TLC poverty of speech: r = -0.38, *p* < 0.001). This is consistent with *d*_0_ indexing the motivational drive to generate verbal output, which is selectively reduced in the context of negative symptoms and alogia. The clinical associations of *d*_0_ are visualized in Figure 2.

**Figure 2:**
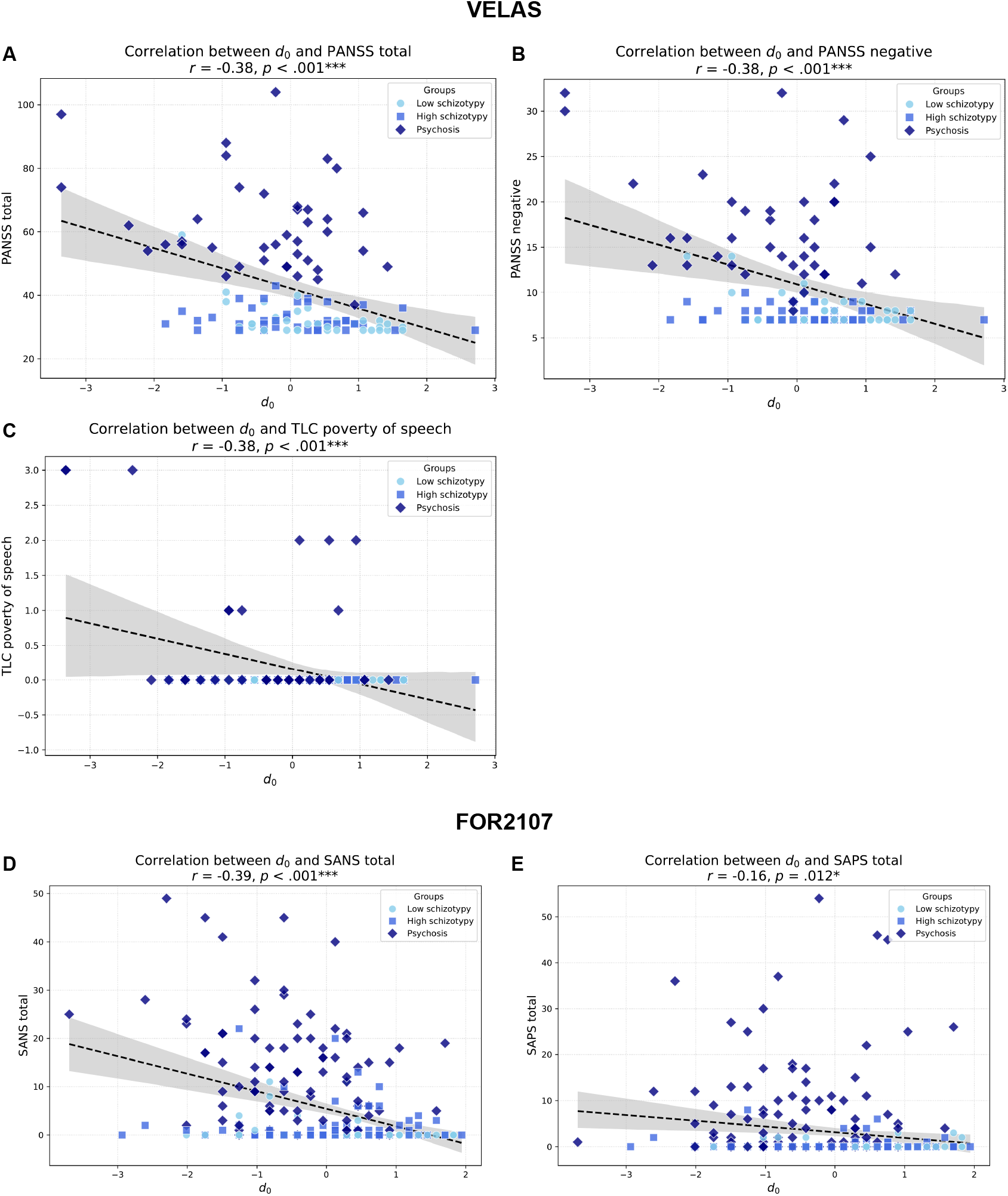
Correlations between initial retrieval drive (*d*_0_) and (A) Positive and Negative Syndrome Scale (PANSS) total score, (B) PANSS negative score, and (C) poverty of speech measured using the Scale for the Assessment of Thought, Language, and Communication (TLC) across all VELAS participants. Correlations between *d*_0_ and (D) Scale for the Assessment of Negative Symptoms (SANS) total score and (E) Scale for the Assessment of Positive Symptoms (SAPS) total score across all FOR2107 participants. Fitted lines are shown for the full samples with 95% confidence intervals..

*α* showed a distinct pattern of associations. In line with the classical notion of loosened associations in psychosis, *α* was negatively associated with positive symptom severity (PANSS positive: r = -0.20, *p* = .031). In addition, *α* was negatively associated with overall FTD during conversational speech (TLC total: r = -0.18, *p* = .051) and disconnectedness (TLC disconnectedness: r = -0.27, *p* = 0.003), indicating that less constrained semantic search co-occurs with greater language disorganization in free speech. Finally, *α* was negatively associated with cognitive flexibility as indexed by the TMT B−A score (r = 0.20, *p* = .033), consistent with the shared executive demands of sustaining and redirecting structured semantic search. These associations are shown in Figure 3.

**Figure 3:**
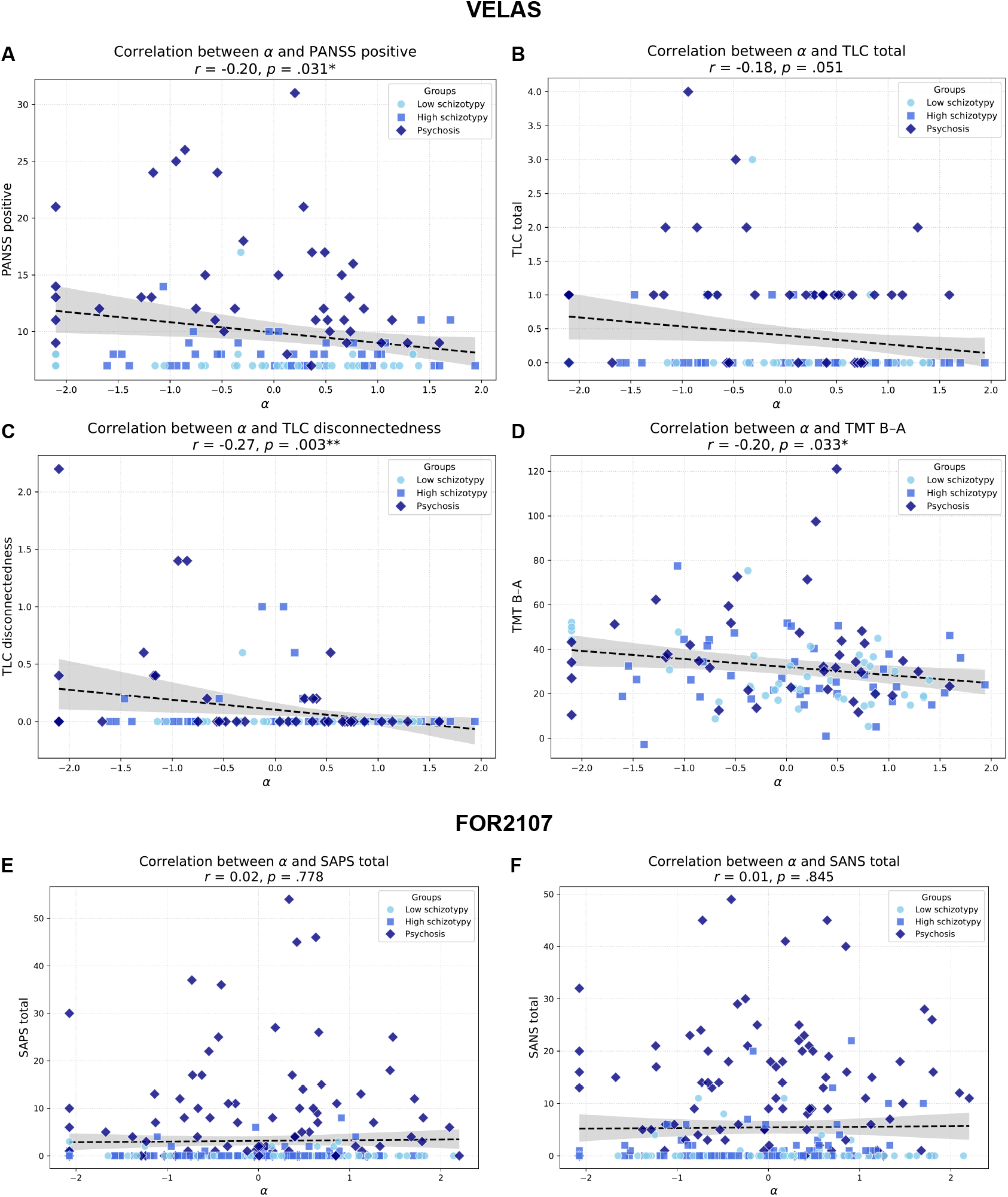
Correlations between semantic search precision (*α*) and (A) Positive and Negative Syndrome Scale (PANSS) positive score, (B) Scale for the Assessment of Thought, Language, and Communication (TLC) total score, (C) disconnectedness measured using the TLC, and (D) cognitive flexibility from the Trail Making Test B minus A score (TMT B–A) across all VELAS participants. Correlations between *α* and (E) Scale for the Assessment of Positive Symptoms (SAPS) total score and (F) Scale for the Assessment of Negative Symptoms (SANS) total score across all FOR2107 participants. Fitted lines are shown for the full samples with 95% confidence intervals.

### Ventral language stream microstructure

The association between *α* and FA of the left ILF was examined both in the full sample, as well as only in the psychosis group, where the hypothesized deficit is most pronounced. Across the full sample, left ILF FA showed a trend-level positive association with *α* (r = 0.18, *p* = .052; Figure 4B), consistent with the predicted direction but not reaching conventional significance. In the psychosis group alone, this association was substantially stronger and significant (r = 0.41, *p* = .009; Figure 4C), indicating that among individuals with early psychosis, those with more preserved white matter microstructure along the ventral semantic pathway also showed more structured semantic search during verbal fluency. This finding connects the computational parameter *α* to a specific neural substrate (the occipito-temporal pathway supporting semantic representation) and is consistent with the broader evidence linking ILF microstructure to semantic processing capacity [29, 31].

**Figure 4:**
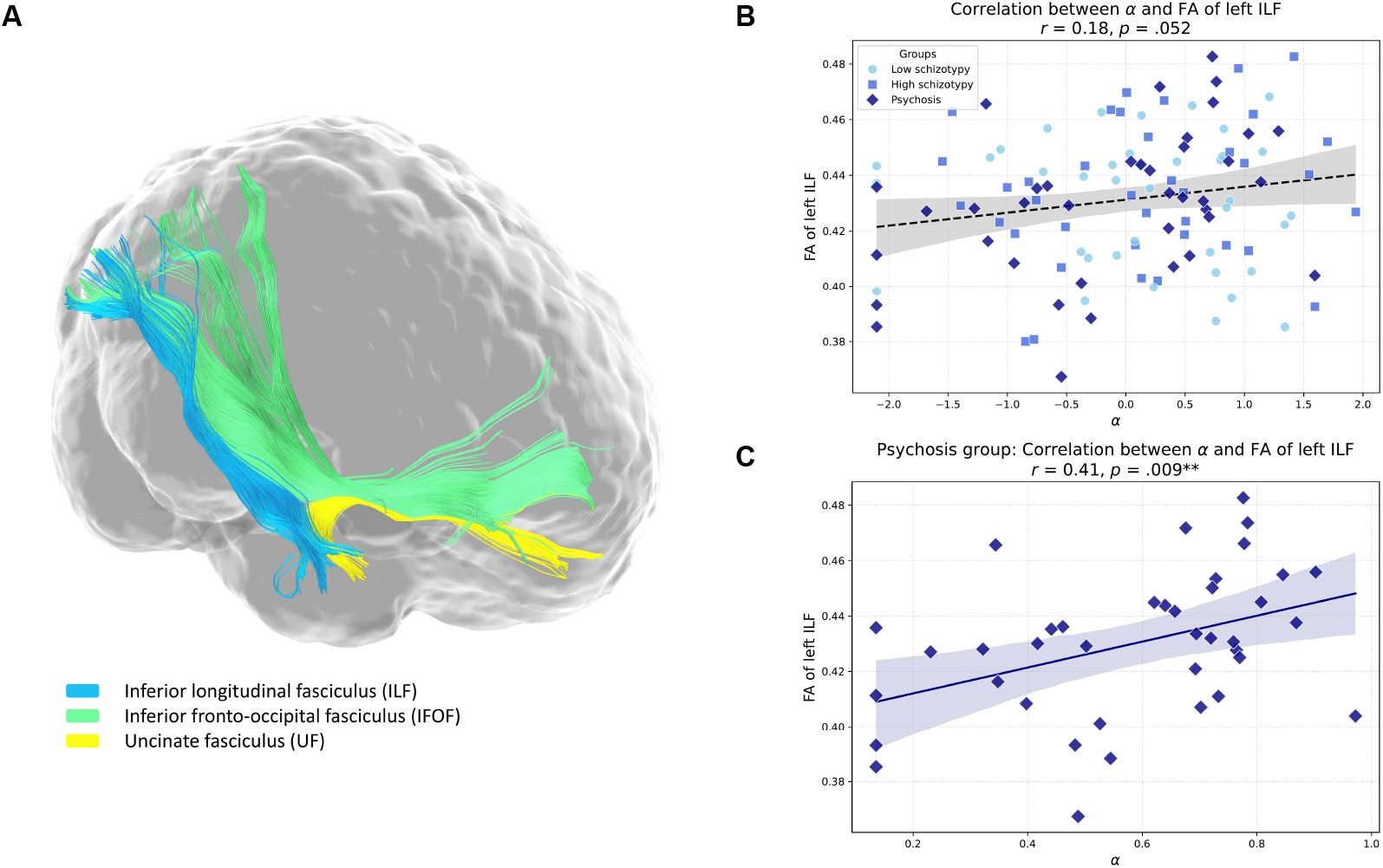
Ventral language stream microstructure and semantic search precision (*α*). (A) Example tractography reconstruction of the ventral language stream in a representative VELAS participant. The ventral stream comprises the inferior longitudinal fasciculus (ILF, blue), the inferior fronto-occipital fasciculus (IFOF, green) and the uncinated fasciculus (UF, yellow), which together link occipital and temporal cortex to frontal regions and support semantic representation. (B) Correlation between *α* and left ILF fractional anisotropy (FA) across all VELAS participants. (C) The same association restricted to the early psychosis group, where the relationship is substantially stronger, linking individual differences in ventral pathway microstructure to individual differences in the precision of semantic search. Fitted lines are shown with 95% confidence intervals.

### Replication sample – FOR2107

#### Sample characteristics

The FOR2107 replication sample comprised 249 participants (*n* = 82 in the low schizotypy, *n* = 84 in the high schizotypy, and *n* = 83 in the schizophrenia group). There were no significant differences between the groups in age (*F*(2, 246) = 0.001, *p* = .999), sex (*χ*^2^(2) = 0.15, *p* = .927), or premorbid IQ (MWT-B IQ: *F*(2, 245) = 1.83, *p* = .162). Compared to the VELAS sample, the FOR2107 sample included significantly older participants (*t* = 10.21, *p* < .001) with higher MTW-B IQ scores (*t* = 12.69, *p* < .001). The distribution of sex did not differ significantly between cohorts (*χ*^2^(1) = 0.525, *p* = .469). Sample characteristics of both samples are shown in Table 1.

#### Clinical measures

The number of items produced in the verbal fluency task differed significantly between groups (*F*(2, 246) = 22.43, *p* < .001). Individuals with schizophrenia produced significantly fewer animals than both the low and high schizotypy groups (*p* < .001 for both comparisons). The results of the clinical contrast replicated those of VELAS: the groups differed significantly in their SAPS (*F*(2, 246) = 44.75, *p* < .001) and SANS scores (*F*(2, 246) = 95.74, *p* < .001), with individuals with schizophrenia showing more severe positive and negative symptoms than individuals with low and high schizotypy (*p* < .001 for each comparison). Descriptive statistics and group comparisons are shown in Table 2.

#### Model parameters

##### Parameter dissociation

The groups differed significantly in their *d*_0_ (*F*(2, 246) = 20.27, *p* < .001). Individuals with psychosis showed significantly lower *d*_0_ than those with either low or high schizotypy (*p* < .001 for each comparison). Again, *d*_0_ showed a strong correlation with the number of items produced (r = 0.94, *p* < 0.001). As in VELAS, *α* did not differ significantly between groups (*F*(2, 246) = 0.53, *p* = .588). Descriptively, *α* was lower in the psychosis group (*M* = 0.61, *SD* = 0.23) than in the low schizotypy group (*M* = 0.62, *SD* = 0.23), but not the high schizotypy group (*M* = 0.58, *SD* = 0.21). *α* correlated significantly with the semantic similarity of consecutive items (r = 0.43, *p* < .001). The correlation between *d*_0_ and *α* was not significant (r = -0.04, *p* = .509), which again underscores that initial retrieval drive and semantic search precision are two dissociable markers of fluency production. Table 2 provides an overview of the descriptive statistics and group comparisons.

##### Clinical associations

Consistent with VELAS, *d*_0_ was negatively associated with negative symptom severity in the FOR2107 sample (SANS total: r = -0.39, *p* < .001), and showed a corresponding negative association with positive symptom burden (SAPS total: r = -0.16, *p* = .012), replicating the pattern of reduced verbal output drive with greater overall psychopathology (Figure 2). Semantic search precision, *α*, showed no significant association with positive symptoms (SAPS total: r = 0.02, *p* = .778), consistent with the general difficulty of detecting symptom-specific effects on *α* in samples where positive symptom variance is limited (Figure 3).

## Discussion

FTD has long resisted mechanistic characterization, in part because the behavioral measures available, clinical ratings and total word counts, do not decompose the underlying process. In this study, we proposed that verbal fluency, one of the most widely administered tasks in clinical neuropsychology, carries separable information about two distinct components of this process: the extent to which motivational drive sustains production (indicated by the initial retrieval drive, *d*_0_), and how strongly retrieval is guided by semantic similarity at each step (indicated by the semantic search precision, *α*). Formalizing this in a computational model, we found evidence of decrease in both components across the psychosis spectrum in two independent samples, yet they dissociated in their clinical correlates as the theory predicted. In the primary VELAS sample, *d*_0_ tracked negative symptom burden and overall psychopathology, as well as the poverty dimension of FTD in conversational speech. *α* tracked positive symptom severity, the disorganization dimension of FTD during conversational speech, and cognitive flexibility. In individuals with early psychosis, *α* additionally correlated positively with the white matter microstructure of the left ILF, suggesting that degraded *α* is grounded in the white matter architecture of the ventral language stream. In the FOR2107 replication sample, the association between *d*_0_ and the negative and overall symptom burden replicated robustly, while the association between *α* and positive symptom severity was flat; most likely because symptom scores in this sample clustered near zero, leaving insufficient variance to detect an effect. *d*_0_ closely tracked list length in both samples, providing evidence that it captures the propensity to sustain verbal output rather than any aspect of the retrieval structure itself.

The correlation of *d*_0_ with negative symptom burden and clinical ratings of alogia in conversational speech is consistent with the broader literature linking reduced verbal output to the poverty dimension of FTD and to the motivational deficits that characterize negative symptoms in schizophrenia [24, 25, 1]. Alogia has long been recognized as one of the most stable and functionally impairing features of schizophrenia-spectrum disorders, and meta-analyses confirm that verbal fluency deficits are among the largest and most replicable cognitive impairments in schizophrenia [47, 48, 17, 18], with evidence that these deficits extend across the psychosis continuum into schizotypy [49, 42, 30]. The present findings suggest that *d*_0_ operationalizes the poverty dimension computationally: it quantifies the resource available to sustain retrieval before stopping, in a way that is continuous, automatically derived, and formally grounded in a generative model of the behavior. Importantly, *d*_0_ and *α* were only modestly correlated in the primary sample and uncorrelated in the replication sample (see Figure S4 in the Supplement), with their clinical associations clearly dissociating. This indicates that reduced drive does not simply reflect a truncated list that happens to have less transition structure. The practical implication is that *d*_0_ may serve as a sensitive and objective index of the poverty dimension of FTD, complementing existing clinical ratings without requiring expert observation.

*α* showed a pattern of associations that was both distinct from *d*_0_ and theoretically coherent. Its correlation with overall FTD and disconnectedness during conversational speech as well as the cosine similarity of consecutive items in the verbal fluency task aligns with a growing body of computational work showing that reduced semantic coherence between consecutive words characterizes the spontaneous speech of people with psychosis and predicts clinical ratings of disorganization [50, 7, 51, 5, 6, 23]. Critically, low *α* in our model corresponded not merely to lower mean similarity but to a flatter selection gradient. This reflects a retrieval process that is structurally less sensitive to the similarity landscape of semantic memory, which is precisely the generative mechanism one would expect to underlie loosened associations as described previously [1]. The association with cognitive flexibility extends this interpretation: structured semantic search requires not only intact semantic representations but also the executive capacity to sustain local search and redirect it when it becomes unproductive, a demand that shares cognitive resources with set-shifting [13, 52, 48, 17, 37]. The finding that *α* correlated with positive symptom severity in the primary VELAS sample but not in FOR2107, where symptom scores clustered near zero, is consistent with the hypothesis that *α* captured a dimension of psychopathology that becomes detectable only when symptom variance is sufficient and cautions against dismissing the association on the basis of the replication null result alone [18, 19, 53].

A structural anchor for the computational parameter *α* is provided by the finding that *α* correlated positively with fractional anisotropy of the left ILF in individuals with early psychosis. The ILF is a major component of the ventral language stream, connecting occipital and anterior temporal cortex and supporting semantic representation [28, 29, 31]. White matter abnormalities along this pathway have previously been reported in schizophrenia-spectrum disorders [54, 26, 55]. The association between *α* and left ILF fractional anisotropy was present as a trend across the full sample and strengthened substantially when restricted to individuals with early psychosis, where both the structural variation and the behavioral deficit were largest. This is consistent with the interpretation that degraded ventral pathway microstructure reduces the precision of the similarity gradient available to guide retrieval, producing the flatter, more random sampling from semantic space that characterizes lower *α*. At the circuit level, this points to parvalbumin interneuron dysfunction reducing lateral inhibitory gain in the temporal semantic cortex [56–58], though this mechanistic account remains a hypothesis to be tested directly in future work. Moreover, together with the cognitive flexibility findings, this paints a coherent picture: *α* reflects a capacity for structured, precision-guided retrieval that depends on both frontal executive resources and the microstructure of the pathway encoding semantic relationships.

A particular strength of the present analysis is that the tracts were reconstructed manually using diffusion-weighted imaging, guided by one of the authors with neurosurgical expertise, rather than relying on automated tractography pipelines prone to systematic errors in regions of complex fiber architecture.

Another strength of our study and a distinctive feature of our model is that stopping is explicitly modeled as a probabilistic process governed by an exponentially decaying drive function, rather than treated as a nuisance or ignored entirely. Most computational accounts of verbal fluency have focused exclusively on the transition structure of lists, leaving the question of when and why speakers stop largely unaddressed [16, 22, 59]. We argue that stopping behavior carries clinically meaningful information, particularly in the context of alogia, and that a model which does not account for it cannot cleanly separate the poverty and disorganization dimensions of FTD.

Several limitations warrant consideration. The analyses are cross-sectional and correlational, precluding causal inference about the relationship between retrieval drive, semantic search precision, clinical symptoms, and white matter microstructure. The model was applied to the semantic verbal fluency task using the category ‘animal’ only; whether the findings generalize to other semantic categories or to letter fluency remains to be established. Word lists of on average twenty to twenty-five items provide limited data per participant, which constrains parameter precision and likely explains the modest effect sizes observed for *α*, whose estimation depends on the transition structure of the list. Future work should aim for extending these sparse data by recording time stamps of retrieval. Furthermore, the two samples differed not only in age and premorbid IQ, but also in their clinical instruments, with PANSS available in VELAS and SANS/SAPS in FOR2107, limiting direct comparability of symptom correlates across samples. Finally, although the mechanistic account linking *α* to parvalbumin interneuron dysfunction and excitation/inhibition balance is theoretically motivated, neither gamma oscillations nor direct markers of inhibitory function were measured, and the neural interpretation therefore remains speculative.

In conclusion, FTD has resisted mechanistic characterization for more than a century, in part because the tools available have described its surface features without decomposing the underlying process. By separating the precision of semantic search from the drive to sustain retrieval, we identified two dissociable computational signatures that map onto the disorganization and poverty dimensions of thought disorder, each with distinct clinical correlates and, in the case of search precision, a specific structural neural substrate. These results open a path toward mechanistically grounded, automatically derived markers of FTD that are sensitive to subclinical variation across the psychosis spectrum and directly interpretable in terms of the cognitive and neural processes they reflect.

## Data Availability

All data produced in the present study are available upon reasonable request to the authors.

## Funding

This project has received funding under the European Union’s Horizon Europe research and innovation programme (grant agreement number 101080251). It is part of the Horizon project TRUSTING. Support was also given by the Swiss National Science Foundation (grant number POZHP1_191938/1), the Brain & Behavior Research Foundation (grant number 28997), and the OPO Foundation (grant number 2020-0075). FS is supported by the German Research Foundation (DFG) through grant STE3301/1-1 (project number 527712970) and by the Von Behring-Röntgen Society (project number 72_0013). FS and TK received funds from the DFG Collaborative Research Centre/Transregio 393 (CRC/TRR 393, project number 521379614).

## Declaration of competing interest

PH has received grants and honoraria from Novartis, Lundbeck, Mepha, Janssen, Boehringer Ingelheim, Neurolite, and OM Pharma outside of this work. No other disclosures were reported.

## Data and code availability

All codes will be made publicly available at https://github.com/homanlab.

## Acknowledgements

We would like to thank Anna Steiner for her valuable contribution to the collection of VELAS data, Nils Lang for his involvement in the initial phases of the study, and Allegra Gasser for her contribution to the manual tractography of the ventral language stream.

## Supplementary information

### Supplementary figures

**Figure S1:**
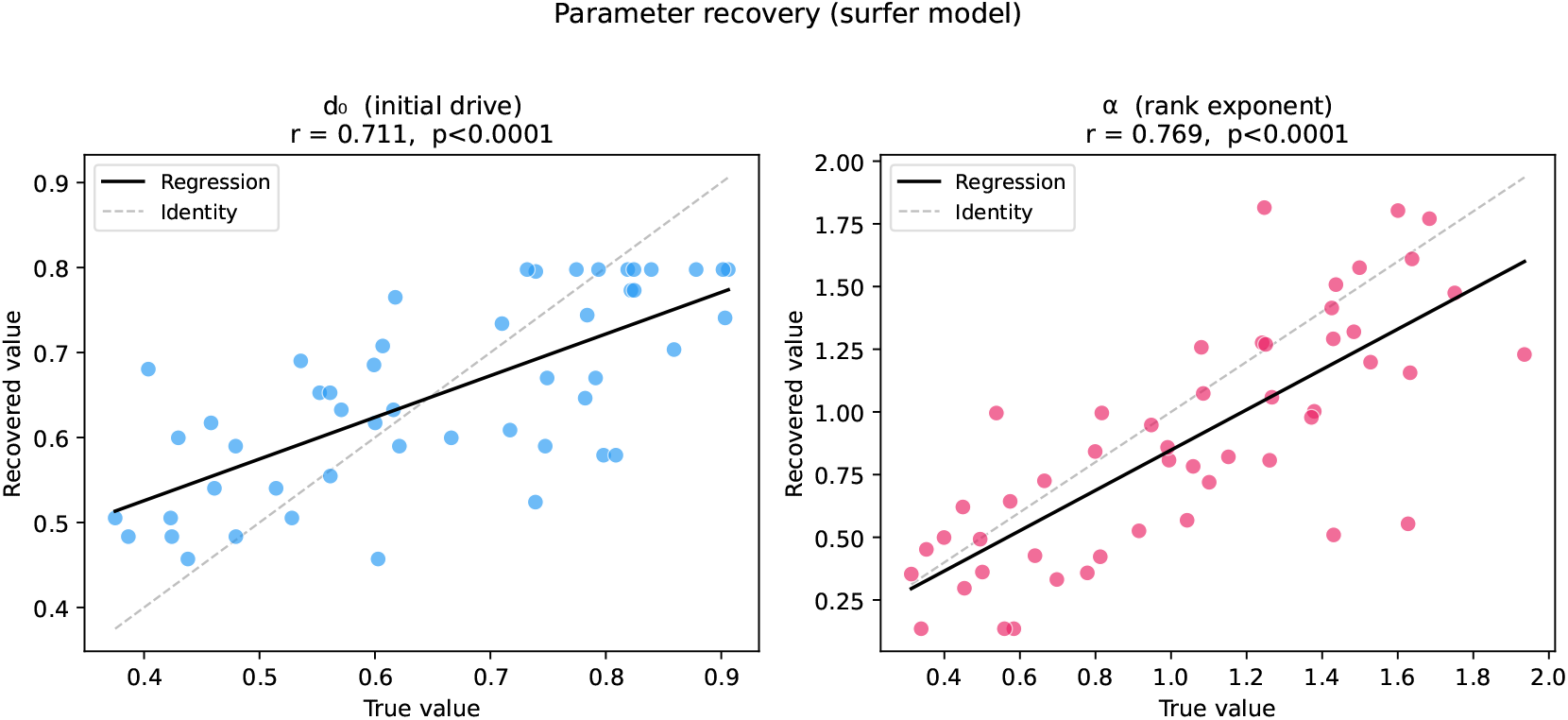
Parameter recovery for the surfer model. Each panel shows the relationship between the true parameter value used to simulate fluency lists and the value recovered by the two-stage EM estimation procedure, across *N* = 50 synthetic participants. True values were drawn from uniform distributions: *d*_0_ ∼ *𝒰* (0.35, 0.92) and *α* ∼ *𝒰* (0.30, 2.00). Fluency lists were simulated from the surfer model using a synthetic block-diagonal similarity matrix (*V* = 60 items, 6 clusters of 10, within-cluster similarity = 0.70, between-cluster similarity = 0.20), and refitted using the same EM procedure applied to the empirical data. Pearson correlations between true and recovered parameters are shown in each panel. Both parameters were recovered with good fidelity, confirming that the two-stage estimation procedure successfully separates the variance attributable to each parameter and that the model is identifiable under conditions approximating the structure of real fluency data.

**Figure S2:**
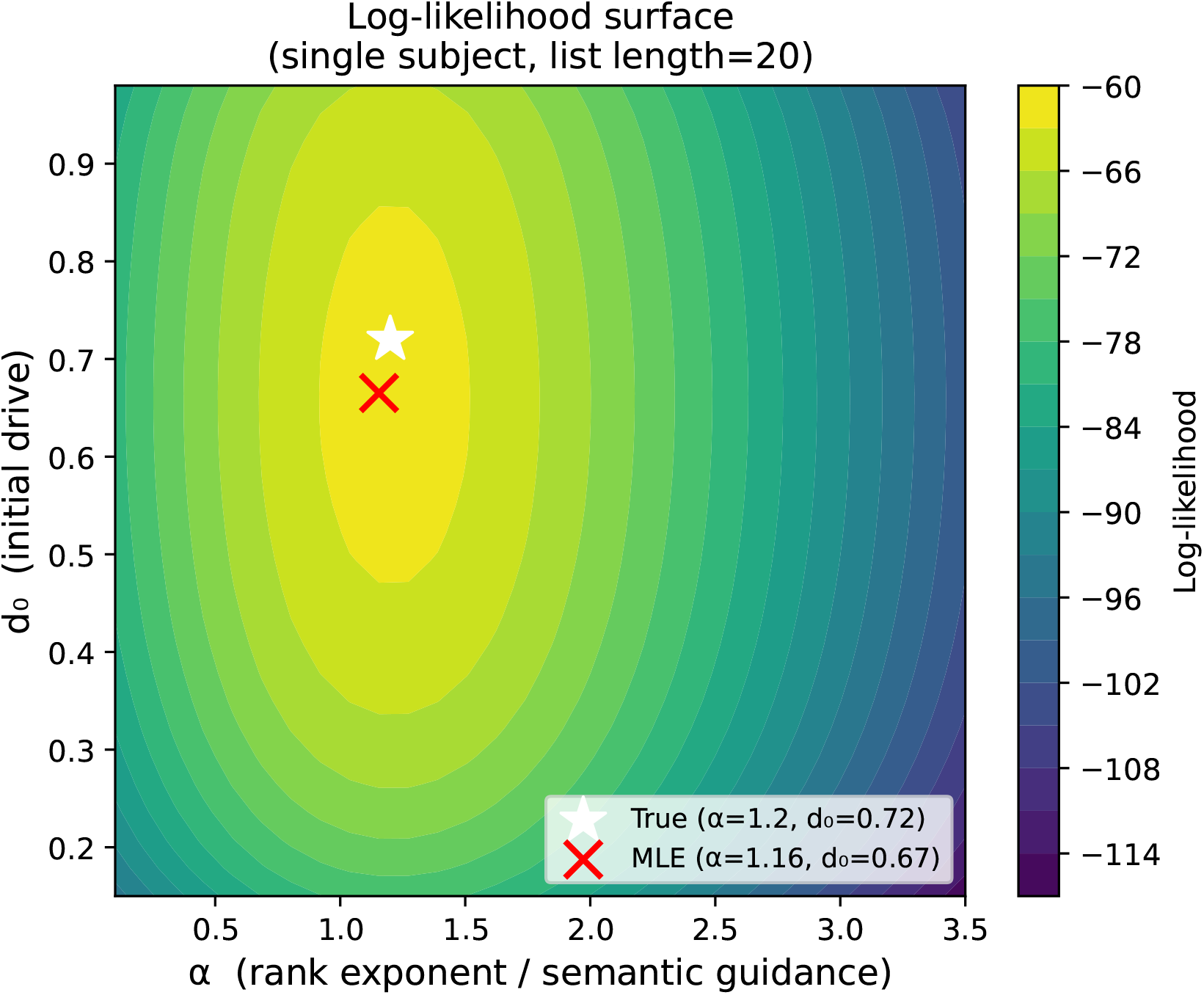
Log-likelihood surface for a representative synthetic participant, shown as a function of *d*_0_ (x-axis) and *α* (y-axis). Warmer colors indicate higher log-likelihood. The surface has a clear, well-defined maximum with no pronounced ridge connecting the two parameter axes, confirming that *d*_0_ and *α* are jointly identifiable and that the two-stage M-step does not introduce a systematic confound between them. This is expected from the model structure: *d*_0_ is identified exclusively by the stopping trajectory of the list, which contributes approximately 3 nats of information per list, while *α* is identified exclusively by the rank-based transition structure, which contributes approximately 115 nats. The large asymmetry in information content motivates the two-stage estimation procedure, which prevents the emission term from dominating the stopping term and destroying identifiability of *d*_0_.

**Figure S3:**
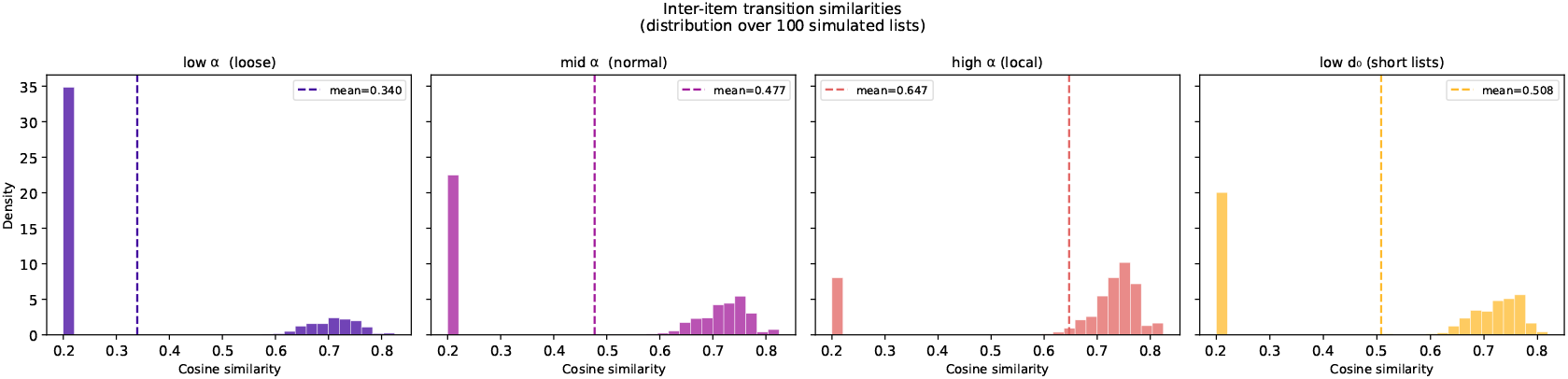
Example fluency lists simulated from the surfer model under contrasting parameter values, illustrating the qualitative difference in retrieval behavior that *α* and *d*_0_ capture. Lists simulated with high *α* show locally coherent, clustered transitions, consecutive items are semantically similar, and the list stays within a narrow region of semantic space for several steps before moving on. Lists simulated with low *α* show a flatter, more random trajectory through semantic space, with consecutive items drawn more uniformly from the available vocabulary regardless of their similarity to the preceding word. Lists simulated with low *d*_0_ are shorter, reflecting earlier stopping, regardless of transition structure. These qualitative patterns correspond directly to the clinical interpretations advanced in the main text: *α* indexes the degree to which semantic similarity guides retrieval at each step, and *d*_0_ indexes the motivational resource sustaining production before stopping.

**Figure S4:**
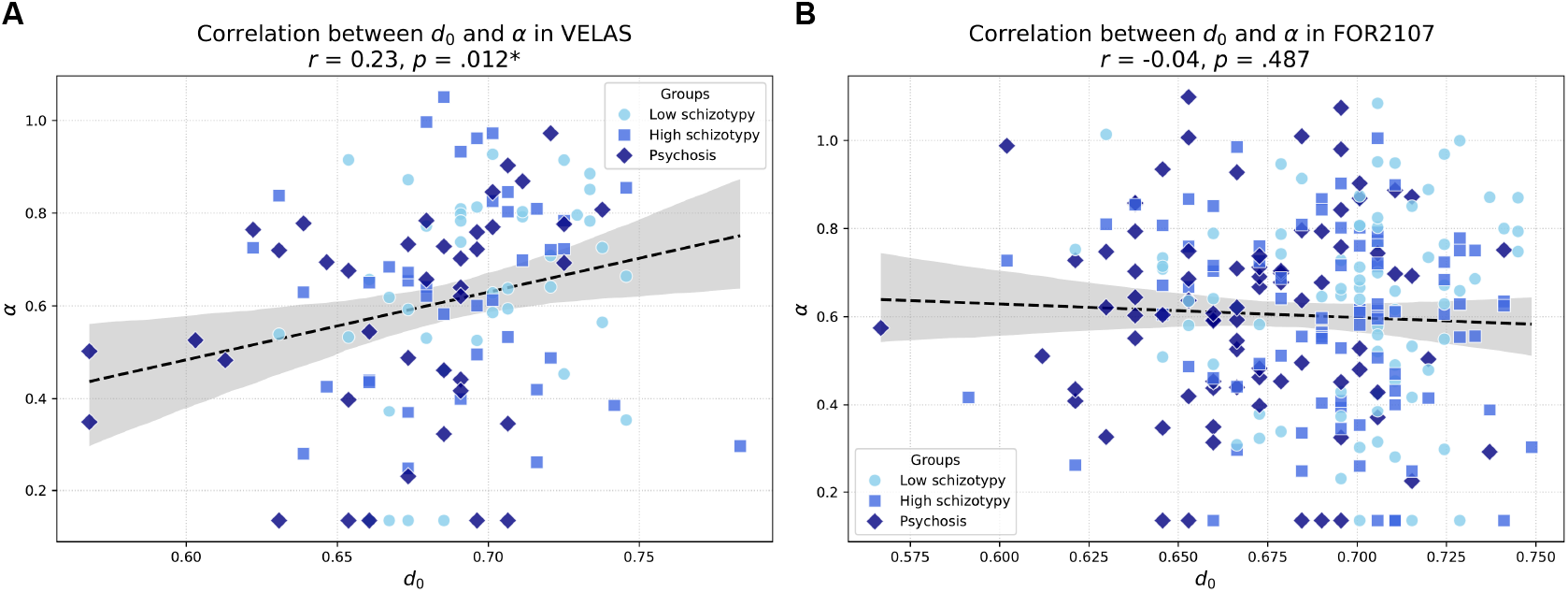
Correlation between initial retrieval drive (*d*_0_) and semantic search precision (*α*) in the (A) VELAS and (B) FOR2107 samples. Fitted lines are shown for the full samples with 95% confidence intervals. The modest correlation in VELAS and absence of correlation in FOR2107 indicate that *d*_0_ and *α* capture dissociable aspects of fluency production, supporting the interpretation that they index distinct dimensions of formal thought disorder. * *p* < .05.

## Notes

### Author Declarations

The Cantonal Ethics Committee Zurich (Kantonale Ethikkommission Zürich; Protocol No. 2021-00347) and the Ethics Committees of the Medical Faculties, University of Marburg (AZ: 07/14) and University of Münster (AZ: 2014-422-b-S) gave ethical approval for this work.

